# Spatial aggregation choice in the era of digital and administrative surveillance data

**DOI:** 10.1101/2021.04.22.21255643

**Authors:** Elizabeth C. Lee, Ali Arab, Vittoria Colizza, Shweta Bansal

## Abstract

**Background:** Traditional disease surveillance is increasingly being complemented by data from non-traditional sources like medical claims, electronic health records, and participatory syndromic data platforms. As non-traditional data are often collected at the individual-level and are convenience samples from a population, choices must be made on the aggregation of these data for epidemiological inference. Our study seeks to understand the influence of spatial aggregation choice on our understanding of disease spread with a case study of influenza-like illness in the United States.

**Methods:** Using U.S. medical claims data from 2002 to 2009, we examined the epidemic source location, onset and peak season timing, and epidemic duration of influenza seasons for data aggregated to the county and state scales. We also compared spatial autocorrelation and tested the relative magnitude of spatial aggregation differences between onset and peak measures of disease burden.

**Results:** We found discrepancies in the inferred epidemic source locations and estimated influenza season onsets and peaks when comparing county and state-level data. Spatial autocorrelation was detected across more expansive geographic ranges during the peak season as compared to the early flu season, and there were greater spatial aggregation differences in early season measures as well.

**Conclusions:** Epidemiological inferences are more sensitive to spatial scale early on during U.S. influenza seasons, when there is greater heterogeneity in timing, intensity, and geographic spread of the epidemics. Users of non-traditional disease surveillance should carefully consider how to extract accurate disease signals from finer-scaled data for early use in disease outbreaks.

## Introduction

Effective disease surveillance systems seek to capture accurate, representative, and timely disease data in the face of complex logistical challenges and limited human resources (1). As these data are typically collected at centralized locations like sentinel healthcare facilities and summarized according political administrative boundaries, there are natural spatial units that may be incorporated into the surveillance system design and reporting. Aggregating surveillance data to administrative boundaries is useful because these units are used in the allocation and distribution of resources and the development of public health guidelines.

While we hope that the spatial and temporal heterogeneity in reported disease surveillance corresponds to the true underlying disease burden, biases in measurement may contribute to inaccurate disease burden estimation. One potential source of bias when working with aggregated surveillance data, often overlooked, stems from choices in the design and aggregation of the reporting data stream itself. In disease ecology, it is well-documented that ecological processes are sensitive to spatial scale, that differences in scale may explain seemingly-conflicting data, and that disease distributions are the result of hierarchical processes that occur on different scales (e.g. 2; 3; 4; 5; 6). Parallel concerns arise in spatial statistics, where the ecological and atomistic fallacies warn against the extension of statistical conclusions from populations to individuals and vice versa (e.g. 7; 8; 9). In epidemiology, methods that account for the hierarchical nature of spatial data have been developed to improve disease mapping and the study of disease dynamics (e.g. 10; 11).

Non-traditional disease data such as digital data streams, syndromic disease reporting, and medical claims were not generated for the purpose of disease surveillance, but they have the potential to provide information relevant to disease tracking in a timely and cost-efficient way across large geographic scales (12; 13; 14; 15; 16; 17). Traditional surveillance systems are designed to meet pre-determined objectives such as routine surveillance or outbreak detection, for a fixed set of syndromes or diseases in a specific population. Non-traditional data are more voluminous and often collected at the individual level, but they often capture a convenience sample limited by user biases. For example, medical claims data captures only individuals with health insurance, while Twitter users with a specific geolocation tag may be younger than the general population in that location. Moreover, the collection of non-traditional disease data is not often designed with attention to logistical reporting constraints. Consequently, epidemiologists and policy makers increasingly have new choices in how to aggregate these records spatially and temporally. Noise and random variability may mask epidemiologically-relevant disease signals in data at finer spatial and temporal scales, and we have limited understanding about how these choices might affect subsequent inference (18; 19; 20). Using U.S. medical claims data for influenza-like illness as a case study, we consider the issue of ‘spatial aggregation choice’ among potentially novel sources of surveillance data. First we characterize influenza season dynamics from 2002-2003 through 2008-2009 across different spatial aggregation scales. We examine defining influenza season features such as the epidemic source location, onset and peak season timing, and epidemic duration with data aggregated to the county and state levels. Finally, we compare spatial autocorrelation for burden between the early and peak influenza seasons, and test the relative magnitude of spatial aggregation differences for seasonal measures related to timing and intensity.

## Methods

### Medical claims data

Weekly visits for influenza-like illness (ILI) and any diagnosis from October 2002 to April 2009 were obtained from a records-level database of U.S. medical claims managed by IMS Health and processed to the county scale, as described elsewhere (21). We also obtained metadata from IMS Health on the percentage of reporting physicians and the estimated effective physician coverage by visit volume (21). Over the years in our study period, our medical claims database represented an average of 24% of visits for any diagnosis from 37% of all health care providers across 95% of U.S. counties during influenza season months (21).

We also aggregated visits for ILI and any diagnosis to the U.S. state- and region-levels, where region boundaries were defined by the groupings of states by the U.S. Department of Health and Human Services.

We performed the same data processing procedure for each county-, state- and region-level time series of ILI per any diagnosis visits (ILI ratio) that has been described elsewhere (21). In brief, ILI intensity is calculated as a detrended ILI ratio that exceeds the epidemic threshold during the flu period from November through March.

### Defining disease burden and spatial aggregation difference

The *intensity* of influenza activity in a given location and time refers to the time series of the detrended ILI ratio. We defined *onset timing* as the number of weeks from week number 40 (first week of October) until the first week in the epidemic period. We defined *peak timing* as the number of weeks from week 40 until the week with the maximum epidemic intensity during the epidemic period. The *epidemic duration* was the number of weeks where the ILI intensity exceeded the epidemic threshold.

We calculated proxies for prevalence during the onset flu season and peak flu season; the *onset intensity* and *peak intensity* metrics were defined as risks relative to the ‘expected’ onset and peak prevalence, respectively. ‘Expected prevalence’ was calculated as the population-weighted mean of the associated intensity measure and it was the same for all counties in a given influenza season. The onset flu season was identified as a 2-3 week flu season period with the greatest exponential growth rate, while the peak flu season was identified as the week with the maximum ILI intensity.

We defined *spatial aggregation difference* as the difference between a given influenza disease burden measure at aggregated spatial scales (i.e., state or region) and the county spatial scale.

### Inferring probable source location

Using seasonal time series of intensity, we identified the top 10% of locations (county or state) with the earliest epidemic onset for each season as potential source locations and calculated the Euclidean distances between the centroids of potential source locations and all other counties. We then used the Pearson correlation coefficient (*H*_*o*_: no difference from zero) between distance to potential source location and onset week to identify probable county or state source locations for a given influenza season (higher correlation coefficient means higher probability of being source location).

### Examining spatial dependence in influenza disease burden

We plotted spatial correlograms to examine the global spatial autocorrelation of the four county-level summary measures of disease burden in the statistical programming language R with the ncf package (22). A two-sided permutation test was performed with 500 permutations to identify correlations that deviated significantly from zero (*H*_*o*_: no difference from zero).

### Comparing spatial aggregation differences across measures and scales

We tested whether spatial aggregation difference was greater among early season or peak season measures of disease burden, and whether state- or region-level aggregations generated greater differences across all measures of disease burden. To compare onset and peak season measures, we paired the spatial aggregation differences for county-season observations across all influenza seasons within our study period for 1) onset timing and peak timing and 2) onset intensity and peak intensity, and tests were performed for both state- and region-level values. To compare differences among state- or region-level aggregations, we paired state-county and region-county differences by county observation for each of the four disease burden measures.

We compared spatial aggregation difference with Bayesian intercept models (effectively, a Bayesian paired t-test) that accounted for county spatial dependence (See SM Methods). The models were implemented with approximate Bayesian inference in R using Integrated Nested Laplace Approximations (INLA) with the INLA package (www.r-inla.org) (23; 24).

Positive estimates mean that 1) spatial aggregation differences for peak timing are greater than those for onset timing, 2) spatial aggregation differences for peak intensity are greater than those for early intensity, or 3) spatial aggregation differences for region and county are greater than those for state and county, and vice versa for negative values. If the 95% credible intervals for *β*_0_ fail to overlap with zero, we interpret that there is a statistically significant difference between the measures contributing to *δ*_*i*_. We used non-informative normal priors for *β*_0_ and non-informative log-gamma priors for the precision term *τ*_*ϕ*_.

## Results

We explore the scales of influenza surveillance using county-level U.S. medical claims data representing 2.5 billion visits from upwards of 120,000 health care providers each year for influenza seasons from 2002-2003 through 2008-2009. There was evident heterogeneity in the intensity and timing of ILI activity between counties and their aggregated state and HHS region scales (Figure 1).

**Figure 1:**
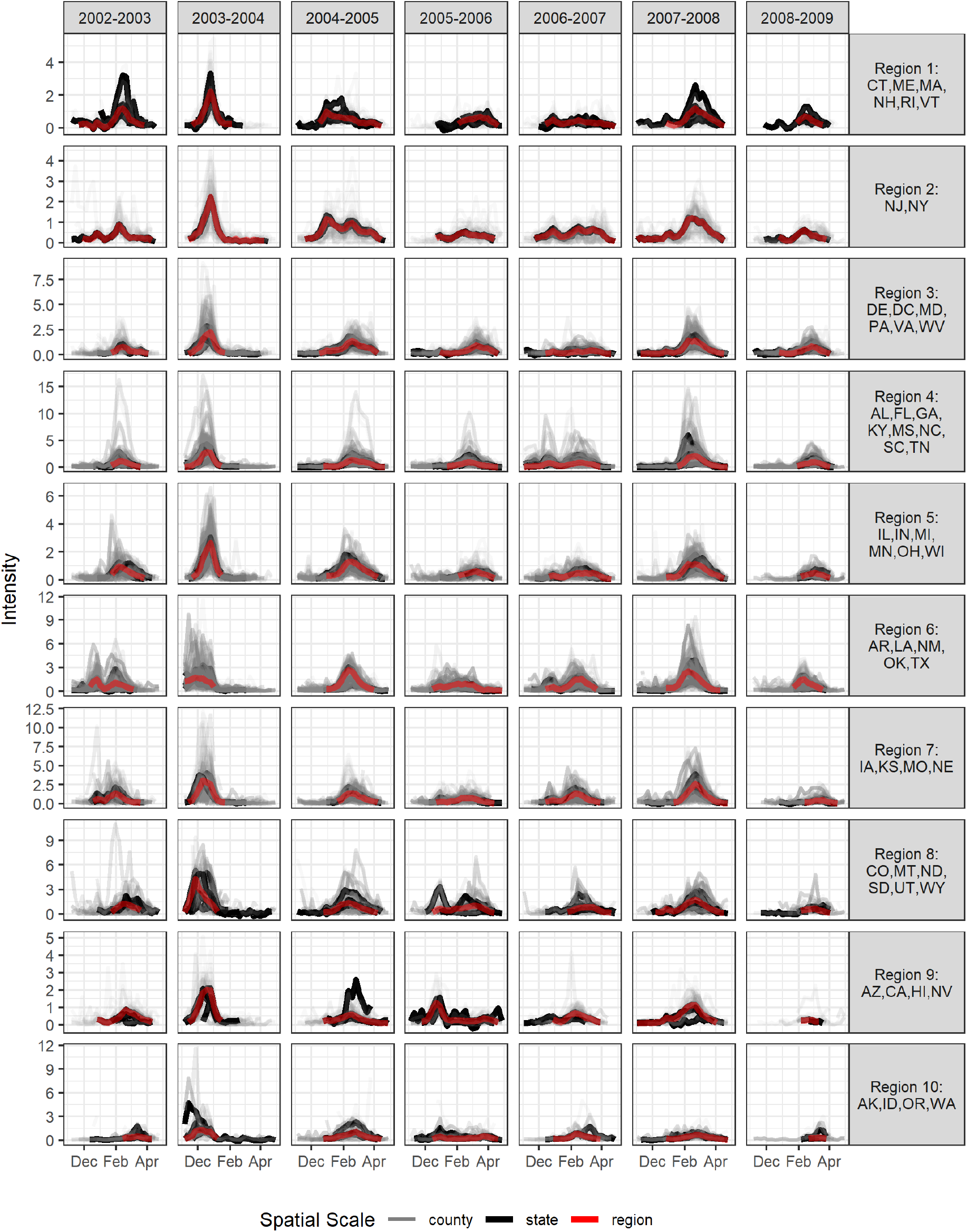
ILI intensity by influenza season from 2002-2003 through 2008-2009 across 10 HHS regions. ILI intensity is displayed for all available counties and states in a given HHS region in different colors (grey for counties, black for states, and red for region).

### Probable epidemic source locations

We inferred the most probable epidemic counties and states independently for each influenza season. We found disagreement in the top two most probable source states and the top 50 most probable source counties (Figure 2). Only in the 2005-2006 influenza season were several probable source counties located within the probable source states (California and Nevada).

**Figure 2:**
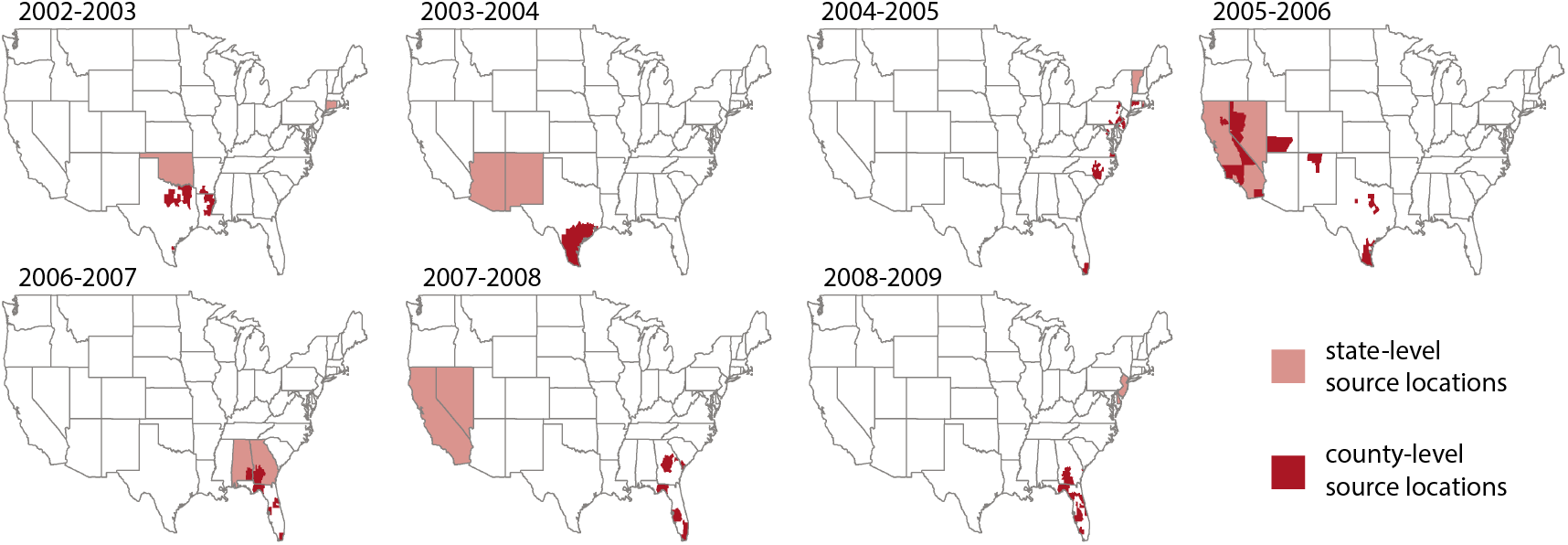
Most probable influenza season U.S. source locations at state and county scales across all influenza seasons. We present the two states (pink) and 50 counties (red) that are the most probable source locations for each influenza season from 2002-2003 through 2008-2009.

### Early season versus peak season

To elucidate the discrepancy between county and state epidemic source locations, we compared the influenza season onset and peak week between county and state scales. While ILI spread was sometimes very rapid, with influenza season onset striking almost all counties within a given state at once, these patterns were not consistent across seasons or states (Figure S1).

State-level flu season onset and peak timing tended to occur after the majority of counties in the state had achieved already achieved those milestones. Across the 2002-2003 through 2008-2009 influenza seasons, a mean of 62% and 70% of state populations had already experienced the onset and peak of the influenza season by the times when the aggregated state-level data achieved its influenza season onset and peak, respectively (Figure 3).

**Figure 3:**
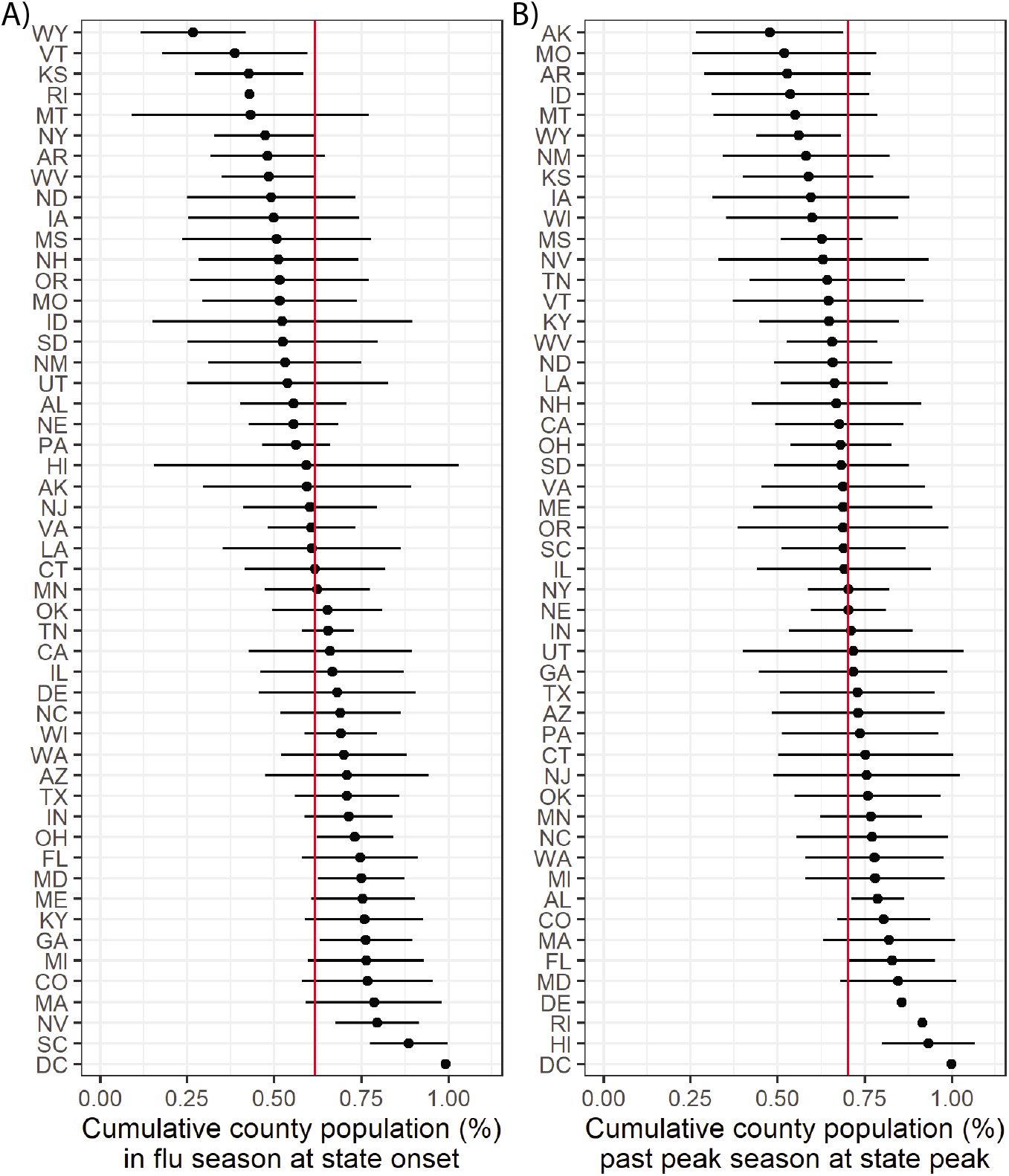
Comparison of county and state influenza season onset and peak timing. We present the cumulative percentage of county populations that have experienced (A) influenza season onset and (B) the influenza season peak by the time that these milestones have been achieved by the aggregated state-level data. For each state abbreviation (rows), the point represents the mean across influenza seasons from 2002-2003 through 2008-2009 while the horizontal line indicates the range of one standard deviation on either side of the mean. The red vertical lines indicate the mean of the mean values across states.

Through visual examination of correlograms, we found that spatial autocorrelation remained present at greater distances for peak measures than early season measures of disease burden, suggesting that seasonal dynamics become more spatially synchronized as the influenza season progresses (Figure 4). Autocorrelation declined to zero at 1177 km and 1359 km for onset and peak timing and at 809 km and 1140 km for onset and peak intensity, respectively.

**Figure 4:**
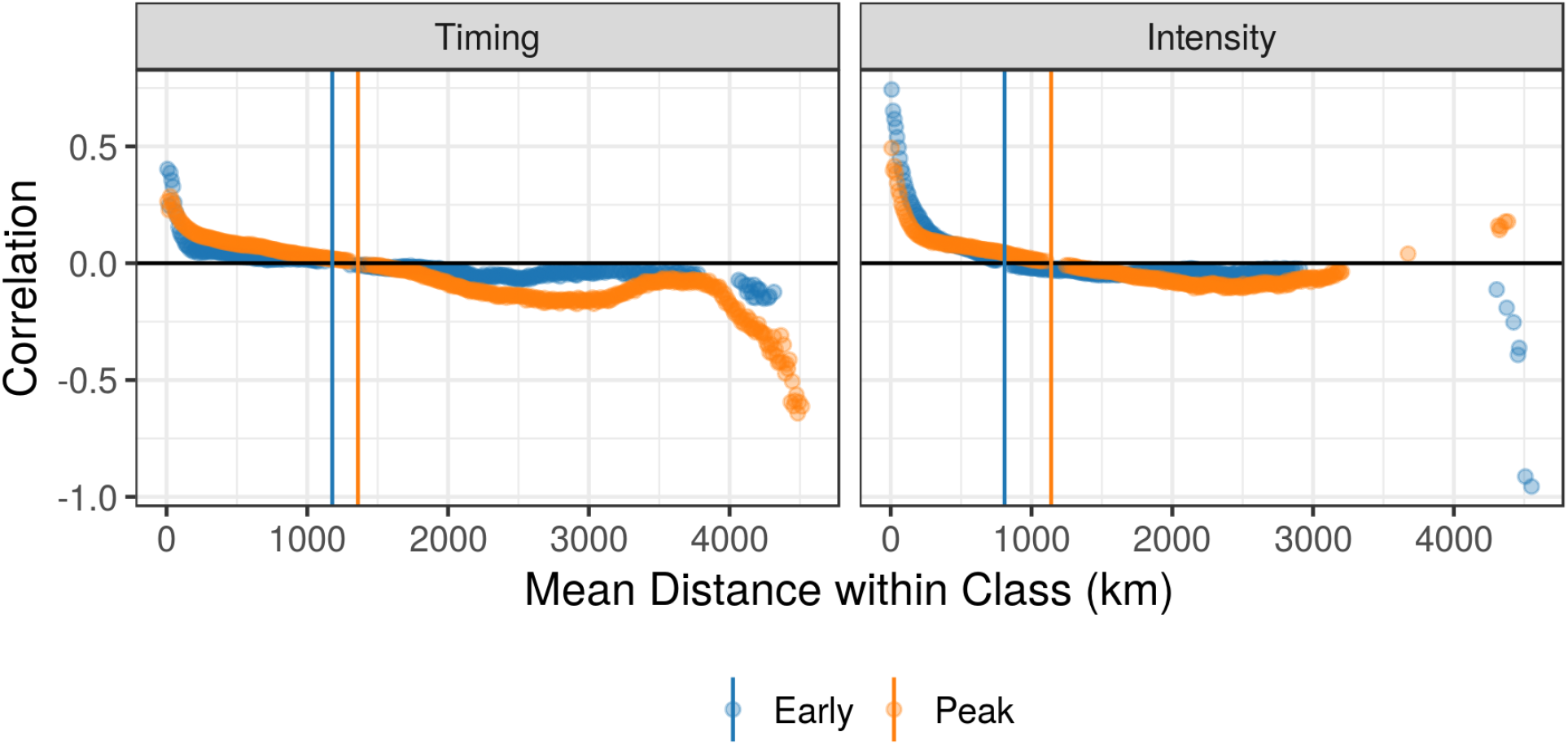
Spatial correlograms for timing and intensity measures across all influenza seasons. We present spatial autocorrelation among counties within specified distance classes for timing measures (left) and intensity measures (right). Early season measures (onset timing and early season intensity) are represented in blue and peak season measures (peak timing and intensity) are in orange. Points are displayed only if the p-value for a two-sided permutation test to evaluate correlation is less than 0.01. Colored vertical lines indicate the mean distance where county measures are no more similar than that expected by chance in a given region.

### Epidemic duration

While county-level epidemics had greater heterogeneity in epidemic duration, often with longer right-skewed tails, epidemic durations were similar across spatial scales (Figure S2). There was greater variability in epidemic duration between influenza seasons than between different spatial scales. Only in the HHS region centered in New York did the distributions in epidemic duration appear to be shifted. However, this region may be particularly subject to discrepancies related to spatial scales because it represents the smallest geographic area in the study region.

### Testing differences in spatial aggregation differences among onset and peak measures of burden

We defined *spatial aggregation difference* as the difference between a given influenza disease burden measure at aggregated spatial scales (i.e., state or region) and the county spatial scale (Figure S3).

We compared spatial aggregation differences between onset and peak timing and between onset and peak intensity using a Bayesian paired t-test. Spatial aggregation differences between state and county measures were greater for onset timing than peak timing and for onset intensity than peak intensity (Table 1). This means that there was greater heterogeneity in the timing and intensity of early season measures than in the peak season measures. Region-county differences were also greater for onset intensity than peak intensity (Table S1).

**Table 1:**
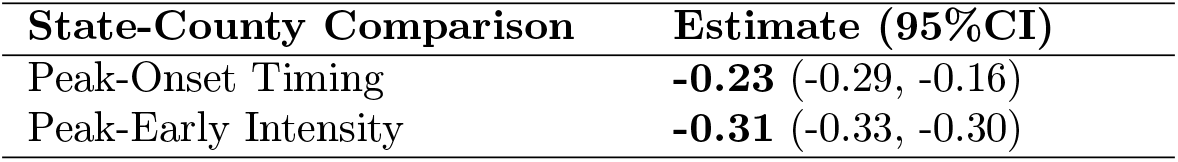
Comparison of state-county spatial aggregation differences between onset and peak season measures. The two negative estimates indicate that peak timing had smaller spatial aggregation differences than onset timing, and that peak intensity had smaller differences than onset intensity. Bolded values denote mean estimates that we interpret to have statistical significance; that is, the 95% credible intervals did not overlap with zero.

## Discussion

In this study, we describe the dynamics and burden of influenza-like illness across the United States from 2002-2003 through 2008-2009 with medical claims data across the county, state, and HHS region spatial scales. We observed substantial heterogeneity in influenza season timing and magnitude across spatial spatial scales and found that analyses performed with county-level and state-level data could provide contradictory results regarding inference on the most probable epidemic source location. State-level timing measures provided delayed information about the onset and peak season timings, and timing-related measures had greater spatial heterogeneity in disease burden and spatial aggregation error than did intensity-related measures.

Our goal was to assess the appropriate usage of potentially novel data streams for influenza surveillance. Administrative health records, social media streams like Twitter, and participatory surveillance systems like Influenzanet and Flu Near You are increasingly available for influenza surveillance, but use of these data for epidemiological analysis is subject to ‘spatial aggregation choice.’ We sought to characterize differences in influenza season features across spatial scales to determine how choice in spatial aggregation may influence subsequent epidemiological inference.

We initially hypothesized that influenza epidemics aggregated to larger spatial scales would have longer epidemic duration and slower spatial spread than county-level data because a state-level or region-level epidemic should represent the set of all lower-level epidemics, which are staggered in time. During our study period, however, state-level onset and peak season timings occurred only after 60-70% of the state’s population had experienced those milestones (as reported by county-level data), while epidemic duration was similar across the county, state, and region scales. The identification of source locations was highly scale-dependent; only in one influenza season were probable source counties located within the probable source state. These results suggest that spatially-aggregated data are less reliable in representing early season dynamics.

We found that the timing and intensity of ILI activity appears to be more heterogeneous in the early season than the peak season, as previously suggested in other settings (25), and heterogeneity is associated with greater spatial aggregation differences (Figure S4-S7). Counties were spatially autocorrelated at greater distances for both peak timing and peak intensity as compared to onset timing and early season intensity, respectively (Figure 4), and spatial aggregation differences were smaller for peak measures than early season measures (Table 1, Table S1). Two factors may contribute to these differences between early and peak season: 1) there are less reliable disease signals during the early flu season, and 2) observations in epidemic onset are asynchronized, but they become more spatially synchronized as the season progresses. Together, these results bolster the hypothesis that seasonal influenza is seeded to many locations and spread primarily through local transmission (26), while prior work suggests that school-holiday-associated contact reductions may play a role in synchronizing influenza outbreaks (27).

Our study suggests that spatial aggregation choice is most critical in early influenza season surveillance (i.e., identifying source locations and early season inference). However, further work should be done to verify the generalizability of our results to different disease syndromes and data sources. Our conclusions about the effects of spatial aggregation may be conflated with the expanding geographic coverage of our medical claims data over time, as well as the stochastic variation in influenza season dynamics itself.

As big data becomes more prevalent and fine-scale targeting and measurement becomes the norm in infectious disease surveillance, spatial aggregation and zoning biases, discrepancies between statistical inference when the boundaries of contiguous spatial units are re-arranged (28; 29), may become regular concerns for epidemiologists. At this juncture, where traditional, administrative, and digital data may be used in disease surveillance, it is critical to develop general methodologies that can extract useful disease signals from fine-scaled data early on in an outbreak (16).

## Data Availability

The medical claims database is not publicly available; they were obtained from IMS Health, now IQVIA, which may be contacted at https://www.iqvia.com/. All model code is available on GitHub at https://github.com/eclee25/flu-SDI-scales.

## Authors’ contributions

ECL and SB conceived the original study. ECL performed the analysis and wrote the first draft of the manuscript. All authors contributed to the study design, advised on analysis, interpreted the data, and edited the manuscript.

## Acknowledgments

This work was supported by the Jayne Koskinas and Ted Giovanis Foundation for Health and Policy [dissertation support grant to ECL], and the Research and Policy for Infectious Disease Dynamics (RAPIDD) program of the Science and Technology Directorate, Department of Homeland Security (DHS), and the Fogarty International Center, National Institutes of Health (NIH).

## Additional Files

**Additional file 1 — Supplementary Material (SM)**

**SM.pdf:** Detailed descriptions about data processing, methodological choices, sensitivity analyses, and supporting evidence. The Supplementary Material (SM) content includes Table S1 and Figures S1 to S7.

