## Supplementary Material for "Spatial aggregation choice in the era of digital and administrative surveillance data"

#### S1 Supplementary Methods

The spatial aggregation differences have spatial dependence and temporal structure with multiple observations per county, thus violating the assumptions of the classical paired t-test. Consequently, comparisons of spatial aggregation difference were assessed with Bayesian intercept models (effectively, a Bayesian paired t-test) that accounted for county spatial dependence (See SM).

$$\delta_i = \beta_0 + \phi_i \tag{1}$$

where $\delta_i$ is defined as the difference between two sets of spatial aggregation differences (e.g., peak versus onset timing, peak versus onset intensity, or region-county versus state-county for a single measure) for county $i$ in a given influenza season. We modeled county spatial dependence $\phi_i$ with an intrinsic conditional autoregressive (ICAR) model, which smooths model predictions by borrowing information from neighbors (30):

$$\phi_i | \boldsymbol{\phi}_{j,-i}, \tau_\phi \sim \text{Normal}(\frac{1}{\xi_i} \sum_{i \sim j} \phi_j, \frac{1}{\xi_i \tau_\phi}), \tag{2}$$

where $\xi_i$ represents the number of neighbors for node $i$, $\boldsymbol{\phi_{j,-i}}$ represents the neighborhood of node $i$, which is composed of neighboring nodes $j$ (neighbors denoted $i \sim j$). The precision parameter is $\tau_\phi$ (Equation 2).

The intercept model was implemented with approximate Bayesian inference in R using Integrated Nested Laplace Approximations (INLA) with the `INLA` package (www.r-inla.org) in order to facilitate the spatial dependence error term (23; 24). If the 95% credible intervals for $\beta_0$ fail to overlap with zero, we interpret that there is a statistically significant difference between the measures contributing to $\delta_i$. We used non-informative normal priors for $\beta_0$ and non-informative log-gamma priors for the precision term $\tau_\phi$.

### S2 Supplementary Results

#### S2.1 Influenza season features across spatial scales

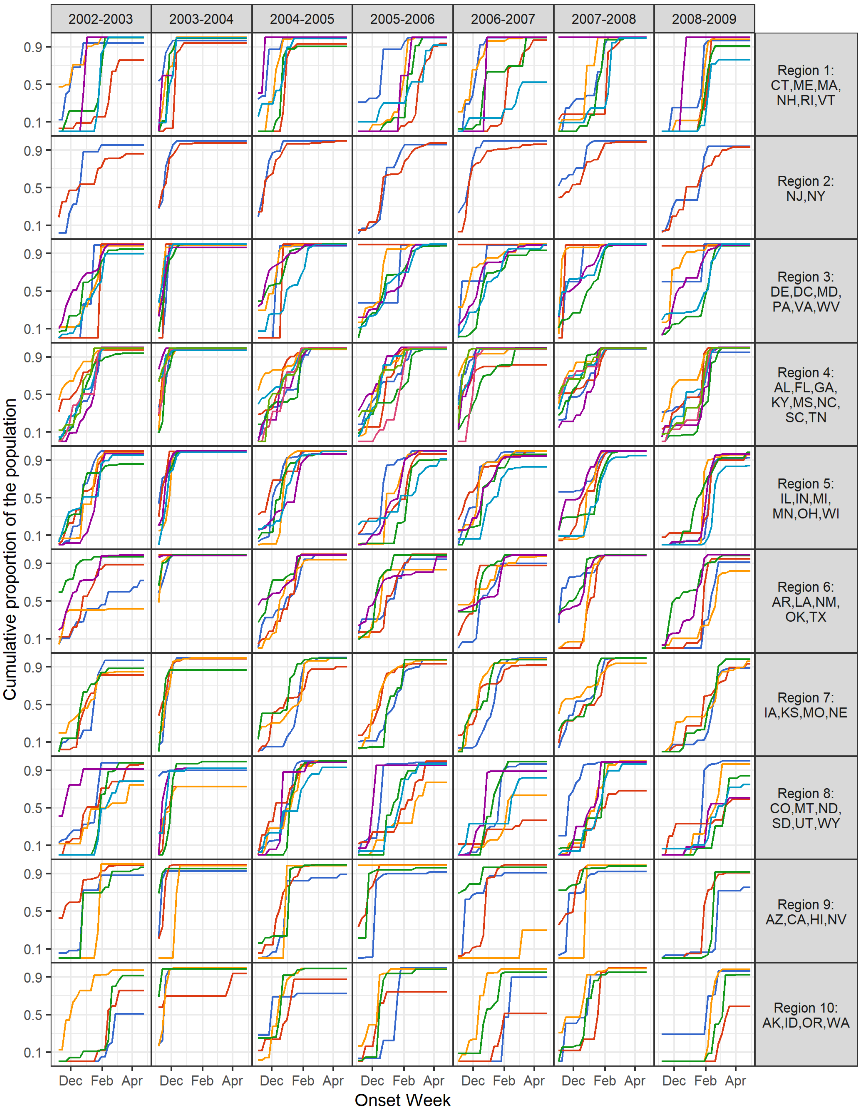


Figure S1: **Summary of onset timing as a function of the cumulative proportion of the population in the influenza season.** We present the cumulative percentage of county populations that have experienced influenza season onset over weeks during the influenza season. Each colored line represents a single state and data for all seasons (columns) are shown grouped by HHS region (rows). Not all counties experience influenza seasons every year, some of the cumulative population proportions remain below one even at the end of the potential influenza season period.

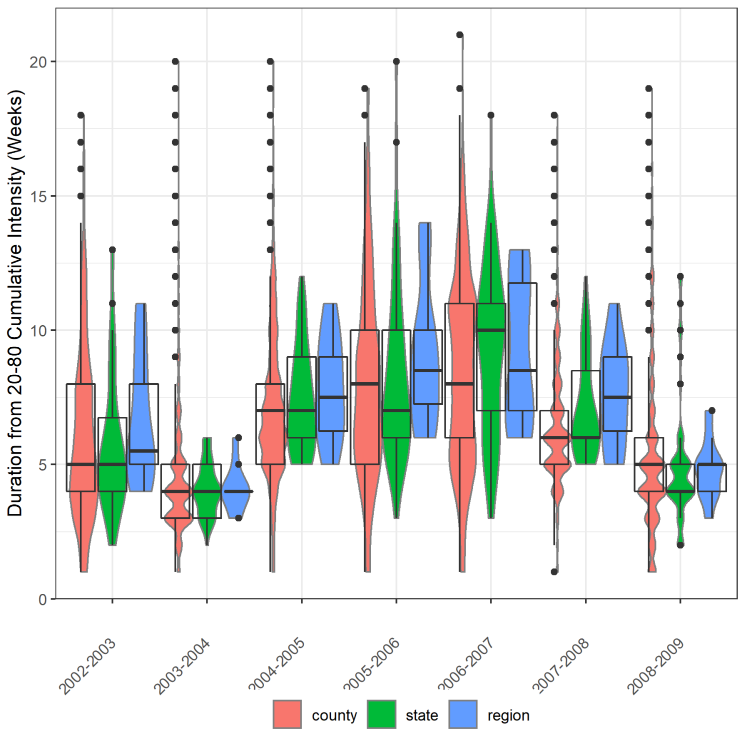


Figure S2: **Epidemic duration by spatial scale.** Epidemic duration was defined as the number of weeks between achieving the 20% and 80% cumulative intensity.

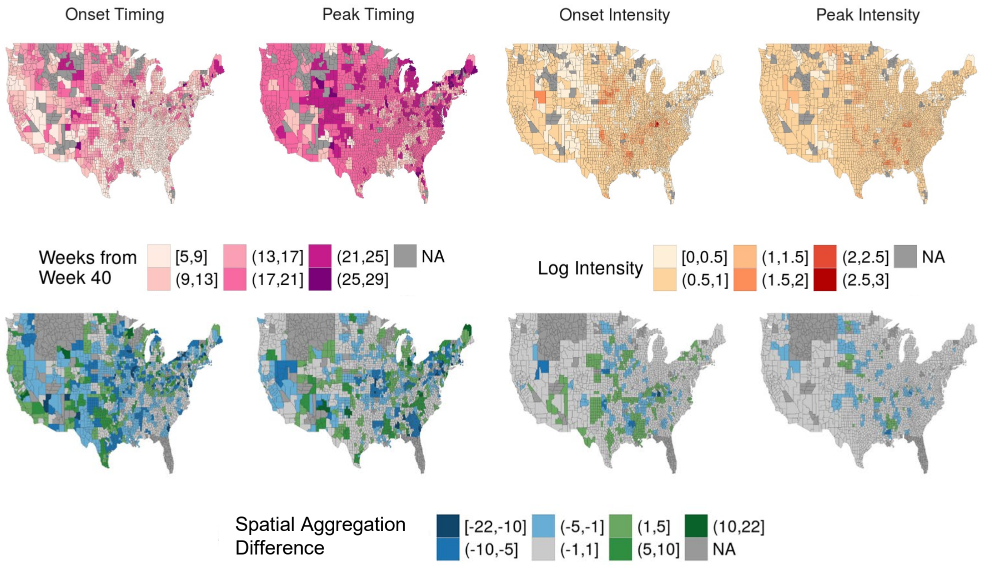


Figure S3: **County-level maps of disease burden and spatial aggregation difference for an example influenza season (2006-2007).** We present county-level disease burden for onset timing, peak timing, onset intensity, and peak intensity (top row) and their associated state-county spatial aggregation differences (bottom row). Timing measures are reported in number of weeks from week 40 while magnitude measures are reported as log intensity. Here, spatial aggregation difference represents the difference between state and county values of disease burden, where negative values (blue) indicate that state-level data underestimated intensity or had earlier timing than county-level data, and vice versa.

Table S1: **Comparison of region-county spatial aggregation differences between onset and peak season measures.** Negative estimates indicate that peak timing had smaller differences than onset timing or that peak intensity had smaller differences than onset intensity. Bolded values denote mean estimates that were statistically significant, where the 95% credible intervals did not overlap with zero.

| **Region-County** | **Estimate (95%CI)** |
|---|---|
| Peak-Onset Timing | **0.07** (0.01, 0.14) |
| Peak-Early Intensity | **-0.48** (-0.50, -0.47) |

#### S2.2 Heterogeneity is associated with greater spatial aggregation differences

We hypothesized that spatial aggregation difference was positively associated with spatial heterogeneity in the disease burden measure of interest. For example, as the county-level variation in peak intensity increases within a given state, we might expect that a state-level estimate of peak intensity would have greater absolute difference. We note that spatial aggregation differences for counties in a given state will not sum to zero; as would be done in public health departments, we process our county and state time series directly from the raw surveillance counts and ILI baselines and epidemic periods are identified independently.

To examine the relation between within-state variation and spatial aggregation difference, we examined the Pearson correlation coefficient between the within-state variance in disease burden and the absolute magnitude of state-county error for each measure and flu season in our study period (28 total comparisons). We found statistically significant positive correlations ranging from 0.34 to 0.95 (p-value less than 0.05) for all but four onset measure comparisons —onset timing for the 2005-06, 2006-07, and 2007-08 seasons and onset intensity for the 2006-07 season (Figure S4, Figure S5, Figure S6, Figure S7). While some results may be unduly influenced by outlier points, we note that the overall pattern indicates a relationship between variation and error.

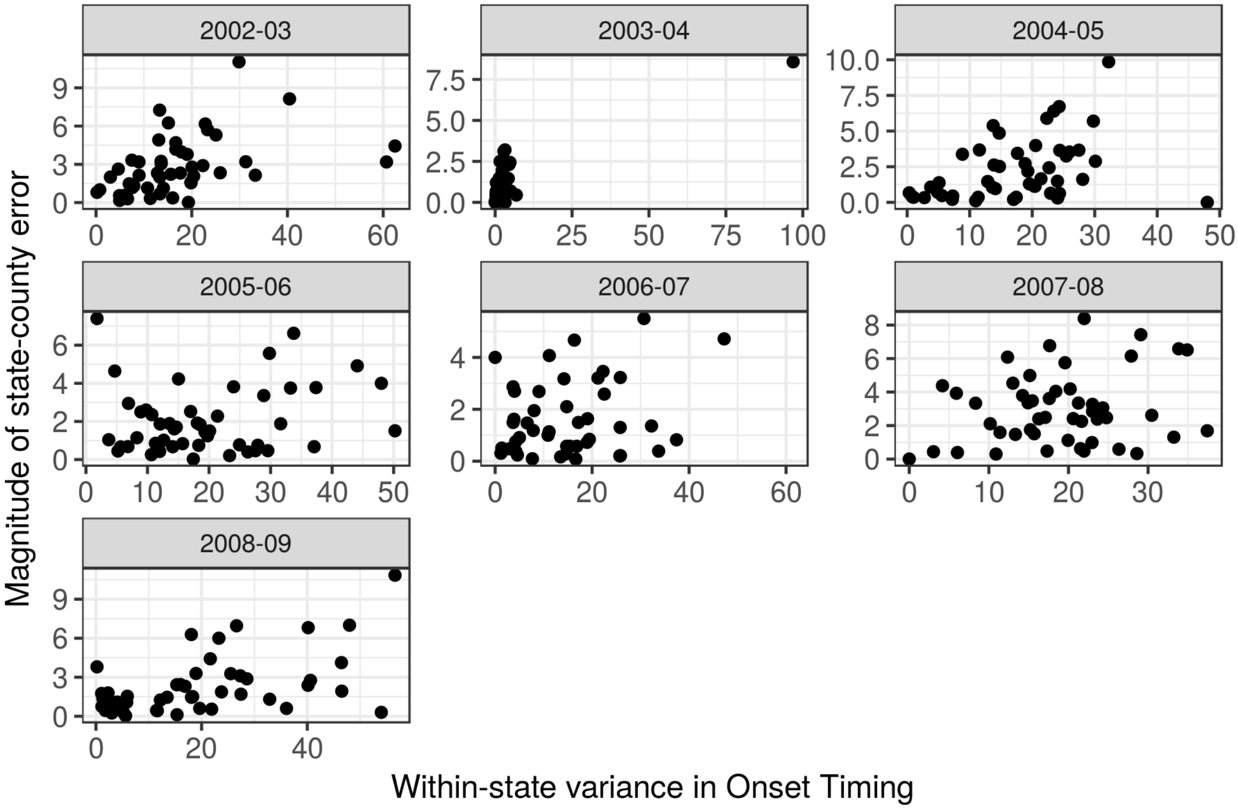


Figure S4: **Scatterplot of spatial aggregation difference and within-state variance in onset timing by influenza season.**

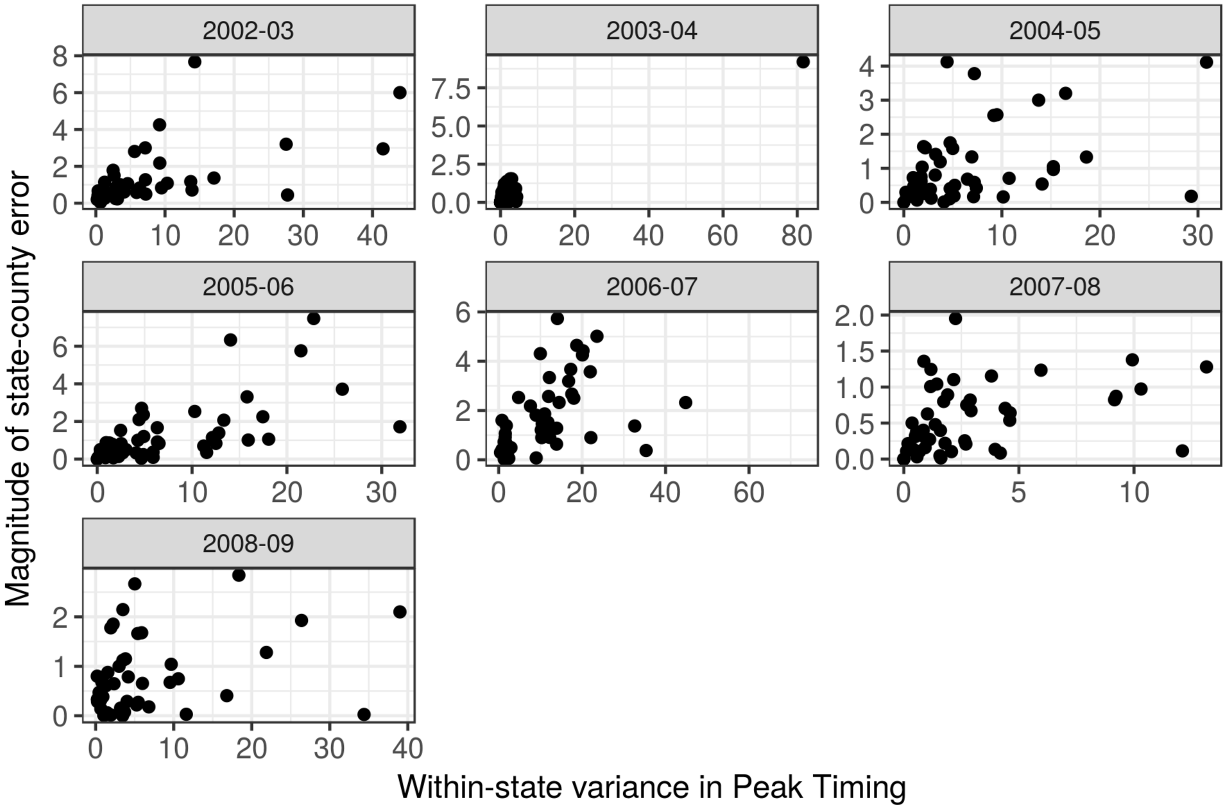


Figure S5: **Scatterplot of spatial aggregation difference and within-state variance in peak season timing by influenza season.**

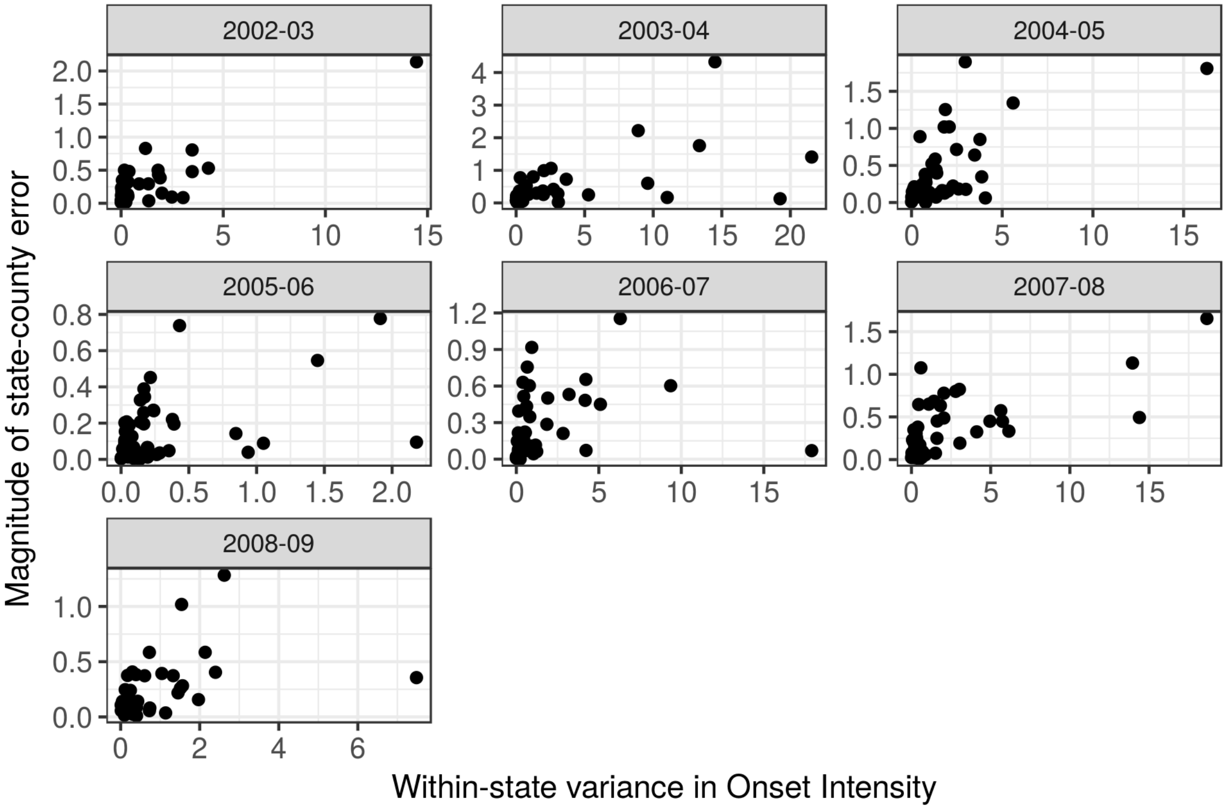


Figure S6: **Scatterplot of spatial aggregation difference and within-state variance in early season intensity by influenza season.**

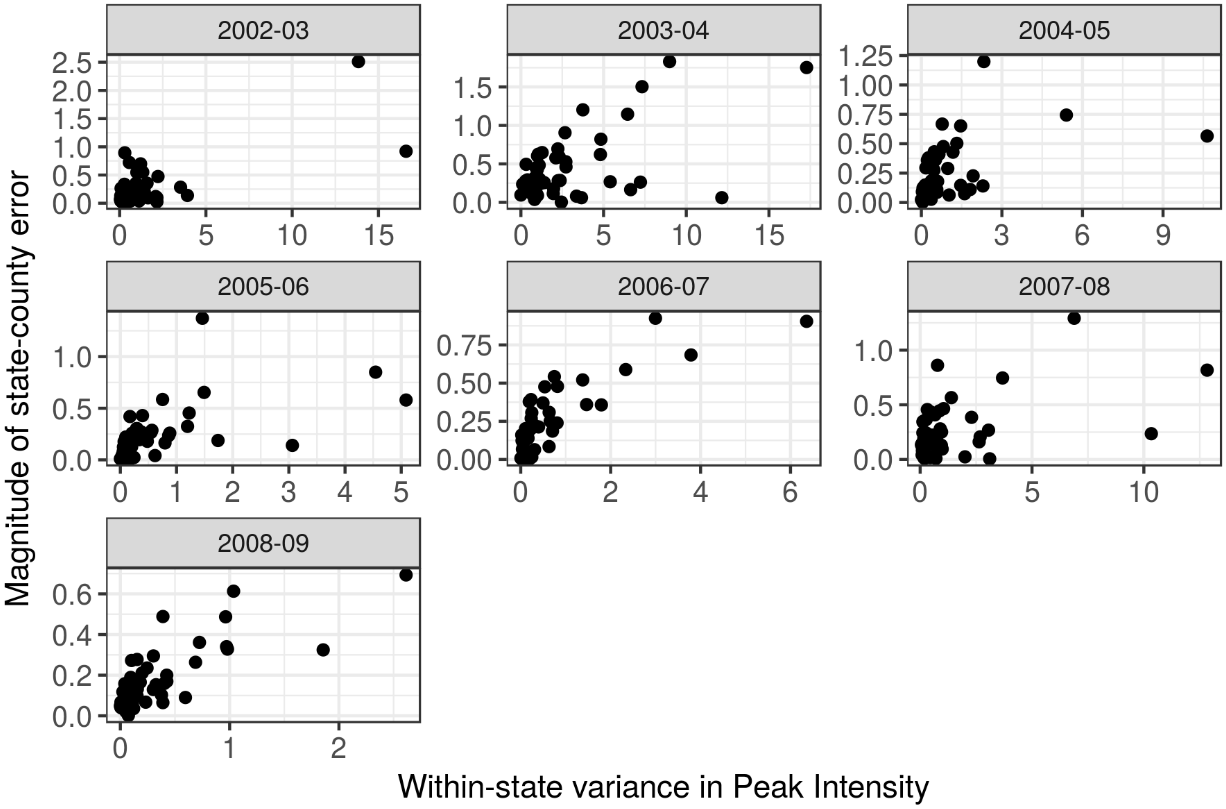


Figure S7: **Scatterplot of spatial aggregation difference and within-state variance in peak season intensity by influenza season.**
